# Restoring Physical Resilience: Consistent Functional and Sarcopenia Gains Across Two Vivifrail Cohorts

**DOI:** 10.64898/2026.10.14.26365508

**Authors:** Javier Fernández-Ortega, Javier Loza-Herbella, Juan A. Rangel-García, Andrés B. Fernández-Revelles

**Affiliations:** Department of Physical Education and Sports, Faculty of Sport Sciences, Sport and Health University Research Institute (iMUDS), University of Granada, Granada, Spain; Santoña Town Council, Cantabria, Spain

**Keywords:** Aged, Frailty, Sarcopenia, Exercise therapy, Physical functional performance, Primary health care

## Abstract

**Objectives:** To evaluate change in physical function and probable sarcopenia following a 36-week community multicomponent exercise programme (Vivifrail model) across two consecutive cycles, and to assess the consistency of this change between cohorts.

**Study design:** Quasi-experimental, single-arm, repeated-measures service evaluation. Ninety-two older adults (2025–2026 cohort; mean age 75.9 years, 88.0% women) completed baseline and post-intervention assessments; an independent 2024–2025 cohort (n = 86) was analysed jointly for replication (n = 178).

**Main outcome measures:** Short Physical Performance Battery (SPPB) score (primary); Timed Up and Go (TUG), handgrip strength, 6-metre walk time and Mini-Mental State Examination (MMSE) (secondary); probable sarcopenia (EWGSOP2), SPPB-based frailty category, and a fall-risk indicator computed identically in both cohorts.

**Results:** SPPB improved by a mean of 1.33 points (Cohen’s d = 0.96; p < .001), with concordant improvement in all four secondary outcomes (all q < .005). 72.8% reached the substantial minimal clinically important difference. Probable sarcopenia fell from 17.4% to 5.4% (p = .006), and frailty category shifted toward higher functional levels (p < .001). Effect sizes were consistent between cohorts for SPPB, handgrip and 6-metre walk; the Time×Cohort interaction was significant for TUG only. In the pooled sample, elevated fall risk fell from 19.7% to 11.2% (p = .002).

**Conclusions:** A 36-week multicomponent exercise programme was associated with clinically meaningful, reproducible improvement in physical function and a reduction in probable sarcopenia across two independent cohorts. The single-arm design precludes causal inference.

## 1. Introduction

Physical resilience, broadly understood as the capacity to maintain or recover functional performance after a physical stressor, is increasingly proposed as a marker of healthy ageing that complements traditional frailty assessment. Frailty itself, a dynamic state of reduced physiological reserve (1), is associated with higher risk of mortality, hospitalisation, falls and disability, although certainty of evidence for several non-mortality associations remains low to moderate (2). Sarcopenia, the age-related loss of muscle mass, strength and physical performance (3), frequently co-occurs with frailty and shares much of its functional expression, particularly reduced handgrip strength and slower gait.

Multicomponent exercise, combining resistance, balance, gait and endurance training, is recommended as the reference strategy for frail and pre-frail older adults because it addresses several determinants of physical resilience simultaneously (4,5). A systematic review and meta-analysis found consistent improvement in the Timed Up and Go (TUG) test, Short Physical Performance Battery (SPPB) score and strength following multicomponent exercise, though evidence for gait speed and long-term maintenance was less conclusive (6). The Vivifrail programme operationalises this evidence as a progressive, community-deployable protocol to prevent functional decline and reduce fall risk (7).

This study builds on two earlier local implementations, a 32-week programme (8) and a 36-week programme in 58 frail older adults (9), which established feasibility but did not examine the consistency of functional change across consecutive delivery cycles or its individual-level clinical meaningfulness. To address this gap, we evaluate a 36-week multicomponent exercise programme delivered in Santoña (Cantabria, Spain) across two consecutive cycles (2024–2025 and 2025–2026), as a municipal public-health service coordinated with primary care. The objectives were: to estimate within-participant change in SPPB and pre-specified secondary outcomes; to quantify the proportion of participants reaching a substantial clinically important improvement, together with change in probable sarcopenia, frailty category and fall risk; to assess whether this change was consistent between the two cohorts; and to explore, within the more recent cohort, whether baseline frailty, age, sex and attendance were associated with the magnitude of change. As a single-arm design without a concurrent control, findings are interpreted as within-participant change during programme participation rather than as a causal effect of the programme.

## 2. Methods

### 2.1. Study design and setting

A prospective, quasi-experimental, single-arm, repeated-measures service evaluation was conducted across two annual 36-week cycles (October–June), with each participant serving as their own reference at baseline (T0) and post-intervention (T1, week 36). A concurrent no-exercise control group was not implemented: the programme is a pre-existing municipal public-health service, coordinated between Santoña Town Council and local primary care, and withholding it from a comparable at-risk group for research purposes was not feasible. As an observational evaluation of routine service delivery in which the intervention was not assigned by the investigators for research purposes, this study was not registered as a clinical trial. This study is reported in accordance with the STROBE (Strengthening the Reporting of Observational Studies in Epidemiology) guidelines.

### 2.2. Participants and eligibility criteria

Participants were recruited through structured referral from primary care; screening was conducted by the sport-science trainer team. Inclusion criteria were age ≥65 years, community-dwelling status, Barthel Index ≥90, TUG ≥10 s, and at least one indicator of functional decline (SPPB ≤10, handgrip <27 kg in men or <16 kg in women (3), or 5STS >15 s). Exclusion criteria were terminal illness or institutionalisation, moderate-to-severe cognitive impairment on the Mini-Mental State Examination (MMSE), unstable cardiac disease or uncontrolled hypertension (systolic >180 or diastolic >110 mmHg), decompensated diabetes or advanced organ failure, and acute or disabling musculoskeletal pathology.

### 2.3. Intervention: multicomponent exercise programme (Vivifrail model)

The intervention followed the Vivifrail programme (7), incorporating strength, balance, gait and flexibility training (10,11). It followed a FITT-VP framework: two supervised group sessions per week (∼66 sessions/cycle), moderate-to-moderately-hard intensity self-regulated using the Borg CR10 scale, 50–60 minutes per session (warm-up, main block, cool-down), 2–3 sets of 8–12 repetitions for resistance work, with individualised progression through resistance, balance complexity and weighted ankle cuffs, delivered by the same qualified exercise-science graduate.

### 2.4. Assessment variables and measurement protocol

The primary outcome was the SPPB total score (12) (balance, gait-speed and chair-rise subtests; 0–12 points; Frail 0–6, Pre-frail 7–9, Robust 10–12 (13,14)); ≥0.5 points was the small and ≥1.0 point the substantial minimal clinically important difference (MCID) (15,16). The feet-together balance subtest showed a complete ceiling effect (100% at maximum) and was excluded from inferential modelling; official point-based scoring (inability = zero points) was used for the SPPB total score and all primary and secondary analyses. For the continuous full-tandem hold and 5STS times specifically, participants unable to complete the task were additionally treated as right-censored (0 s / 60 s) for a supplementary Tobit sensitivity analysis (17).

Secondary outcomes were handgrip strength (Jamar dynamometer, American Society of Hand Therapists protocol (18), EWGSOP2 cut-offs (3)), TUG [20], 6-metre habitual gait-speed time, and the MMSE (19), the latter also used as an exclusion screen. Age, sex and attendance (sessions completed/offered) were recorded for all participants.

### 2.5. Fall risk classification

Elevated fall risk was defined as a TUG time ≥13.5 s or a gait speed <0.80 m/s (6-metre time >7.50 s) (20,21), applied identically from raw TUG and gait-speed data in both cohorts.

### 2.6. Ethics and sex/gender analysis

This study was conducted in accordance with the Declaration of Helsinki as a service evaluation and quality-improvement analysis of an established municipal public-health exercise programme referred from primary care. In accordance with national regulations for non-experimental healthcare service audits (Spanish Law 14/2007 on Biomedical Research), formal Institutional Review Board approval was not required, as the evaluation used secondary analysis of fully anonymised functional data collected during routine service delivery; no intervention was assigned for research purposes, and the study was accordingly not registered as a clinical trial. Informed consent was obtained from all participants. Sex was recorded as a binary biological attribute (male/female) from administrative records, gender identity was not separately assessed.

### 2.7. Statistical analysis

Analyses used R 4.5.1 (tidyverse, rstatix, lme4/lmerTest, emmeans, effectsize, coin, ggpubr, corrplot, survival, splines, ggalluvial packages). Continuous variables are mean ± SD; categorical variables are n (%). SPPB was the sole primary confirmatory outcome; TUG, handgrip, 6-metre walk and MMSE were secondary outcomes; the 4-metre gait and 5STS subtests were analysed descriptively as SPPB components rather than independent confirmatory outcomes.

Paired pre–post comparisons used the Wilcoxon signed-rank test (paired t-test for reference), with repeated-measures Cohen’s d and 95% CI (formulae in Supplementary Methods). The MCID responder proportion was calculated with bootstrap 95% CI (2,000 resamples); a multidomain responder met the MCID in at least two of SPPB, TUG and handgrip. Frailty-category and sarcopenia transitions were evaluated with McNemar’s test; the three-category frailty matrix additionally used a zero-cell-robust McNemar–Bowker test, presented alongside an alluvial diagram. Within the 2025–2026 cohort, linear mixed models tested Time×subgroup interactions (sex, age group, baseline frailty) with a random intercept per participant, and independent-samples comparisons by sex are reported for baseline characteristics.

The primary dose-response analysis used linear regression of SPPB change on attendance, adjusted for baseline SPPB, age and sex; restricted cubic splines and a pooled, cohort-adjusted repeat were exploratory analyses (Supplementary Methods). Change-score correlations are reported descriptively in the Supplementary Material. Cross-cohort replication used linear mixed models on the four shared core outcomes (SPPB, TUG, handgrip, 6-metre walk), with time, cohort, age and a Time×Cohort interaction as fixed effects and a random participant intercept; for SPPB, the between-cohort difference in change was additionally compared against a pre-specified equivalence margin of ±0.5 points. Baseline cross-cohort homogeneity used the Wilcoxon rank-sum test. Benjamini–Hochberg correction was applied specifically to the four secondary outcomes; α = .05 for all two-tailed tests.

## 3. Results

### 3.1. Participant flow and baseline characteristics

A total of n = 92 older adults enrolled and completed both baseline and post-intervention assessments, constituting the full analysis set (Supplementary Figure S1); n = 42 met the per-protocol criterion of ≥80% attendance. Baseline characteristics, overall and by sex, are shown in Table 1; no sex difference reached significance for any baseline characteristic. At enrolment, 17.4% (n = 16) met EWGSOP2 criteria for probable sarcopenia, and the sample spanned all three frailty categories (Frail 4.3%, Pre-frail 23.9%, Robust 71.7%).

**Table 1.** Baseline characteristics of the full analysis set, overall and by sex.

| Characteristic | All (n = 92) | Women (n = 81) | Men (n = 11) | p |
| --- | --- | --- | --- | --- |
| Age, years, mean (SD) | 75.9 (6.8) | 76.1 (6.9) | 74.5 (6.1) | .47 |
| SPPB total score, mean (SD) | 10.11 (1.90) | 10.09 (1.92) | 10.27 (1.79) | .77 |
| **Frailty category, n (%)** |  |  |  |  |
| Frail (0–6 pts) | 4 (4.3) | 4 (4.9) | 0 (0.0) |  |
| Pre-frail (7–9 pts) | 22 (23.9) | 19 (23.5) | 3 (27.3) |  |
| Robust (10–12 pts) | 66 (71.7) | 58 (71.6) | 8 (72.7) | .81<sup>a</sup> |
| Probable sarcopenia (EWGSOP2), n (%) | 16 (17.4) | 13 (16.0) | 3 (27.3) | .38 |

<sup>a</sup>Fisher's exact test, three-category frailty distribution (women vs men); all other p-values from Welch's t-test (continuous) or Fisher's exact test (categorical).

### 3.2. Primary and secondary functional outcomes

SPPB improved from a mean of 10.11 to 11.43 points (+1.33; dᵣ = 0.96, 95% CI 0.71–1.21; p < .001; Figure 1), the largest effect of any measure. Among the secondary outcomes, TUG improved (dᵣ = 0.59; p < .001), handgrip increased (dᵣ = 0.51; p < .001), 6-metre walk time improved (dᵣ = 0.25; p = .005), and MMSE improved (dᵣ = 0.51; p < .001); all four remained significant after Benjamini–Hochberg correction (q < .005). At the SPPB-component level, 4-metre gait (dᵣ = 0.33) and 5STS time (dᵣ = 0.47) also improved (both p < .001). A supplementary Tobit sensitivity analysis indicated the naive approach to censored tandem-hold times underestimated the true improvement (+1.78 s) relative to the Tobit estimate (+6.08 s); effect magnitudes were broadly similar in the per-protocol subset (Supplementary Table S1).

**Figure 1.**
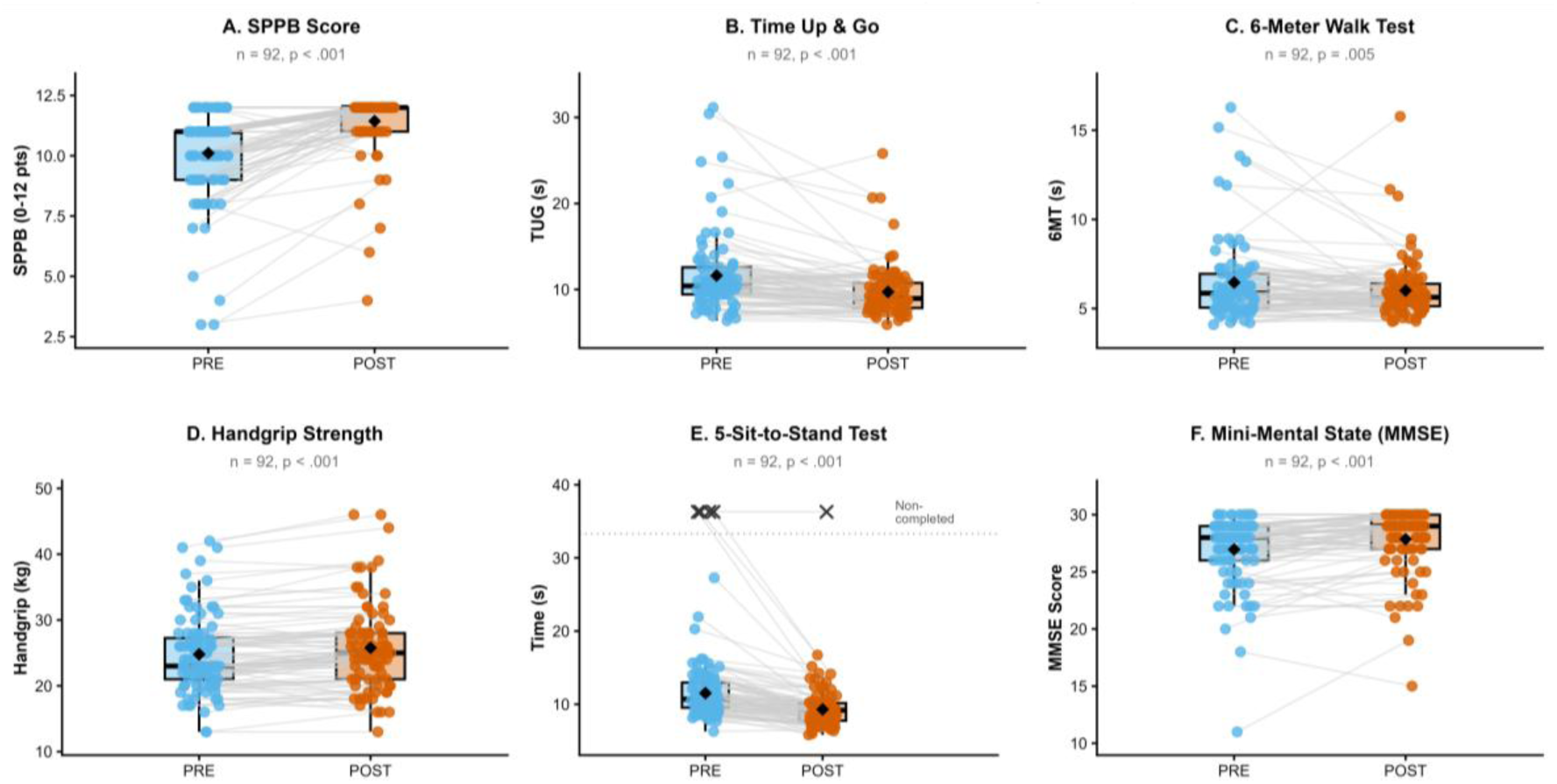
Pre-to-post functional and cognitive changes in the full analysis set (n = 92). (A) SPPB total score (primary outcome), (B) TUG time, (C) 6-metre walk time, (D) handgrip strength, (E) 5STS time (SPPB component), (F) MMSE score.

### 3.3. Reversal of probable sarcopenia and frailty status transitions

Probable sarcopenia prevalence fell from 17.4% to 5.4% (McNemar p = .006; Figure 2), a 12.0-percentage-point reduction. SPPB-based frailty category shifted toward higher functional levels (zero-cell-robust McNemar–Bowker test: p < .001; Figure 3); a robust-versus-non-robust sensitivity analysis confirmed a marked movement into the robust category (p < .001).

**Figure 2.**
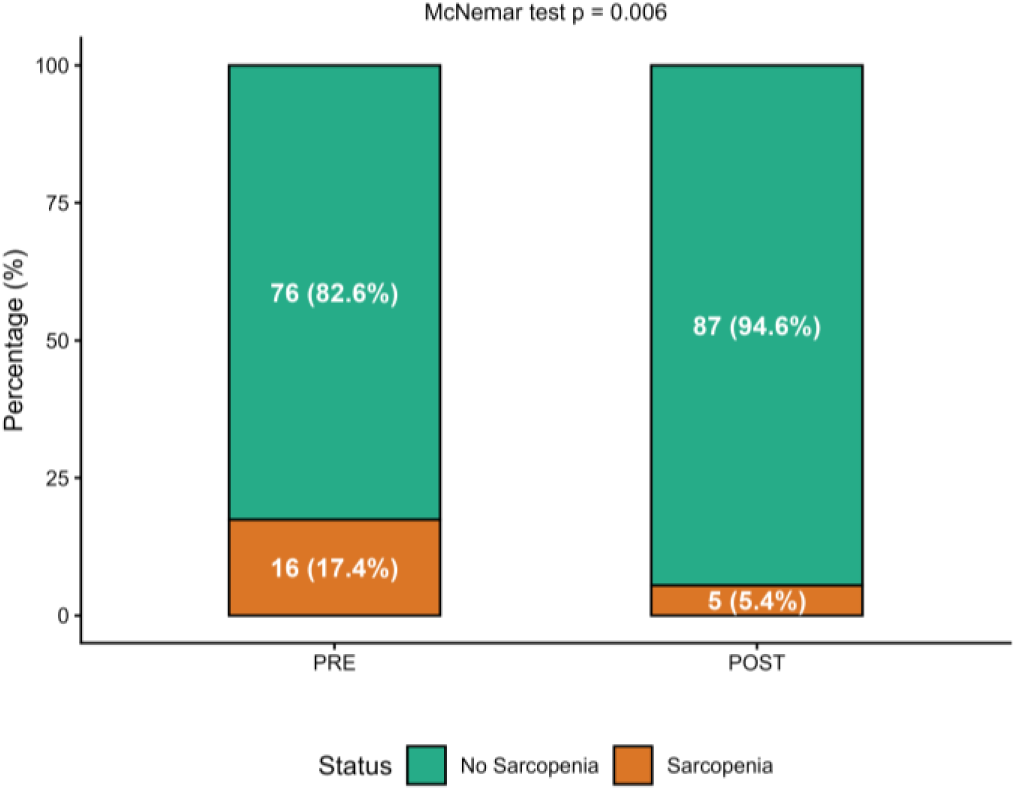
Probable sarcopenia (EWGSOP2) prevalence before and after the programme, full analysis set (n = 92).

**Figure 3.**
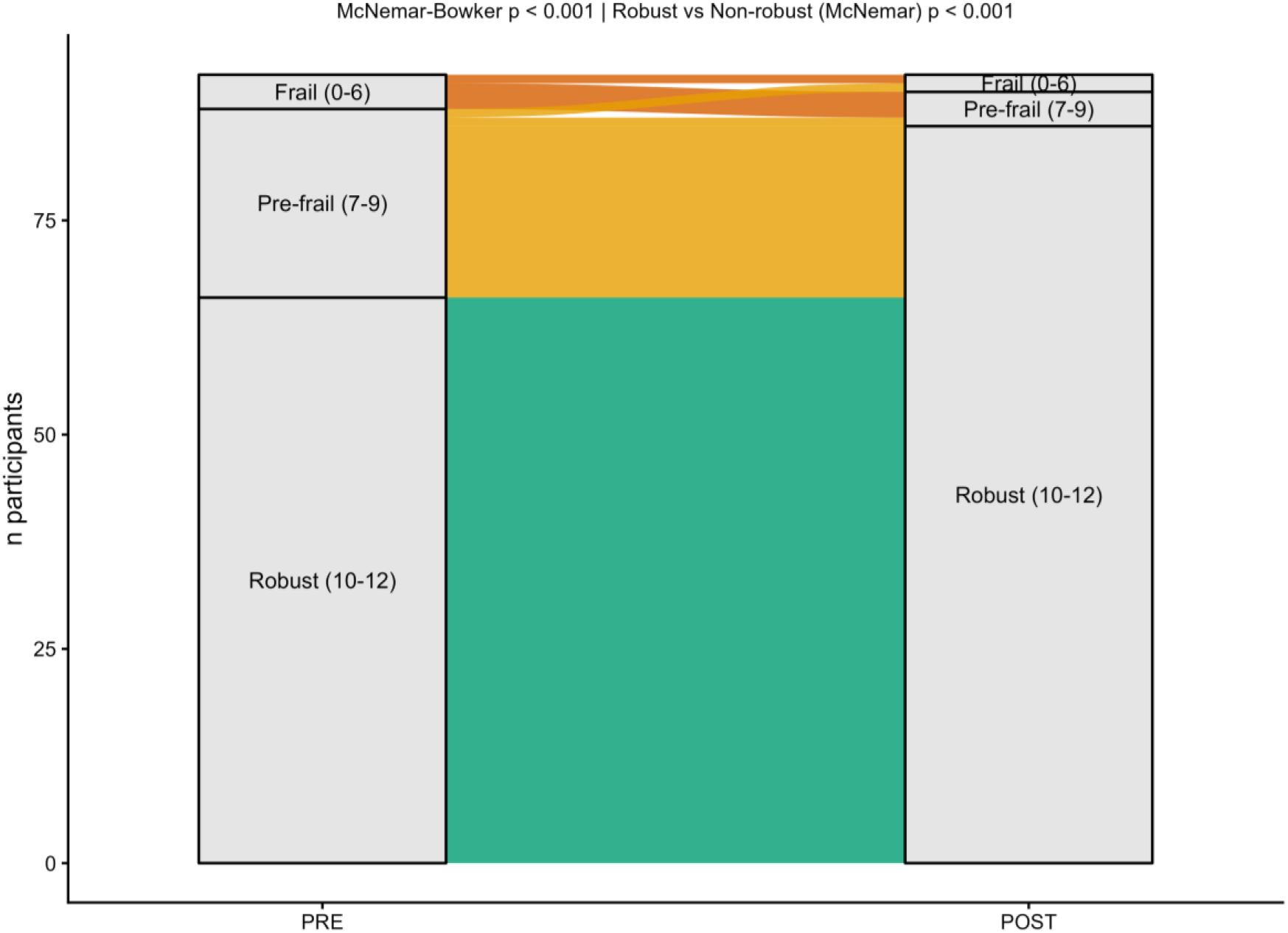
Frailty category transition (SPPB-based) from baseline to week 36, full analysis set (n = 92). Frail (0–6), Pre-frail (7–9), Robust (10–12).

### 3.4. Individual clinical responsiveness and subgroup consistency

Against the substantial MCID threshold (Figure 4), 72.8% of the full analysis set (n = 67/92; bootstrap 95% CI 64.1–81.5%) were clinical responders, 22.8% showed no change, and 4.3% (n = 4) declined. This rate was consistent across adherence, age and sex strata (Supplementary Table S2). A multidomain responder analysis (MCID in ≥2 of SPPB, TUG, handgrip) identified 42.4% (n = 39/92) as multidomain responders. Linear mixed models found no statistically significant Time×subgroup moderation of the SPPB time-effect by sex, age group or baseline frailty (all p > .05).

**Figure 4.**
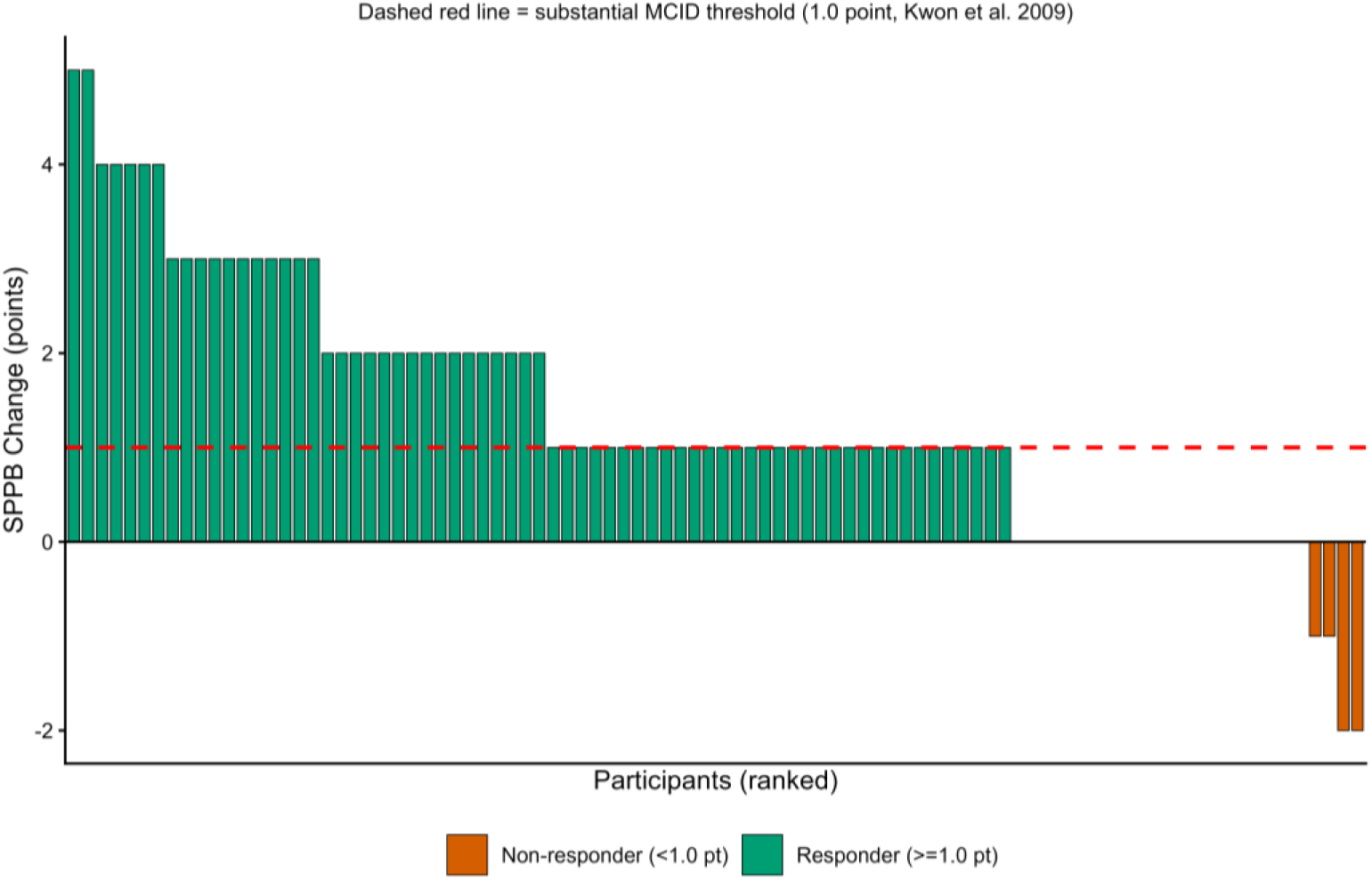
Waterfall plot of individual SPPB change (POST−PRE), full analysis set (n = 92), ranked and coloured by responder status relative to the substantial MCID.

### 3.5. Dose-response and inter-domain covariation

A multiple linear regression adjusted for baseline SPPB, age and sex found no independent association between attendance and SPPB change (p = .81); baseline SPPB was the dominant predictor (p < .001). Supplementary restricted cubic spline models, within-cohort and pooled, were consistent with this linear result (Supplementary Figures S2 and S3). Inter-domain change-score correlations (Supplementary Figure S4), the full effect-size profile (Supplementary Figure S5) and subgroup responder rates (Supplementary Figure S6) are also reported descriptively in the Supplementary Material.

### 3.6. Pooled and replication cohort analyses

Baseline homogeneity between cohorts was confirmed for age, SPPB and adherence (all p > .05). Cohort-specific effect sizes across the four shared core outcomes showed broadly overlapping confidence intervals (Figure 5): SPPB (2024–2025 dᵣ = 0.87 vs. 2025–2026 dᵣ = 0.96), handgrip (0.54 vs. 0.51) and 6-metre walk (0.31 vs. 0.25). The Time×Cohort interaction was non-significant for SPPB (p = .067), handgrip (p = .51) and 6-metre walk (p = .90), but significant for TUG (p = .001), which was larger in 2025–2026 (dᵣ = 0.59) than in 2024–2025 (dᵣ = 0.26). For SPPB, the between-cohort difference in change was −0.35 points (95% CI −0.72 to 0.02); because this interval does not lie entirely within the pre-specified ±0.5-point margin, strict equivalence cannot be claimed for SPPB, although the difference is modest and the omnibus interaction was non-significant.

**Figure 5.**
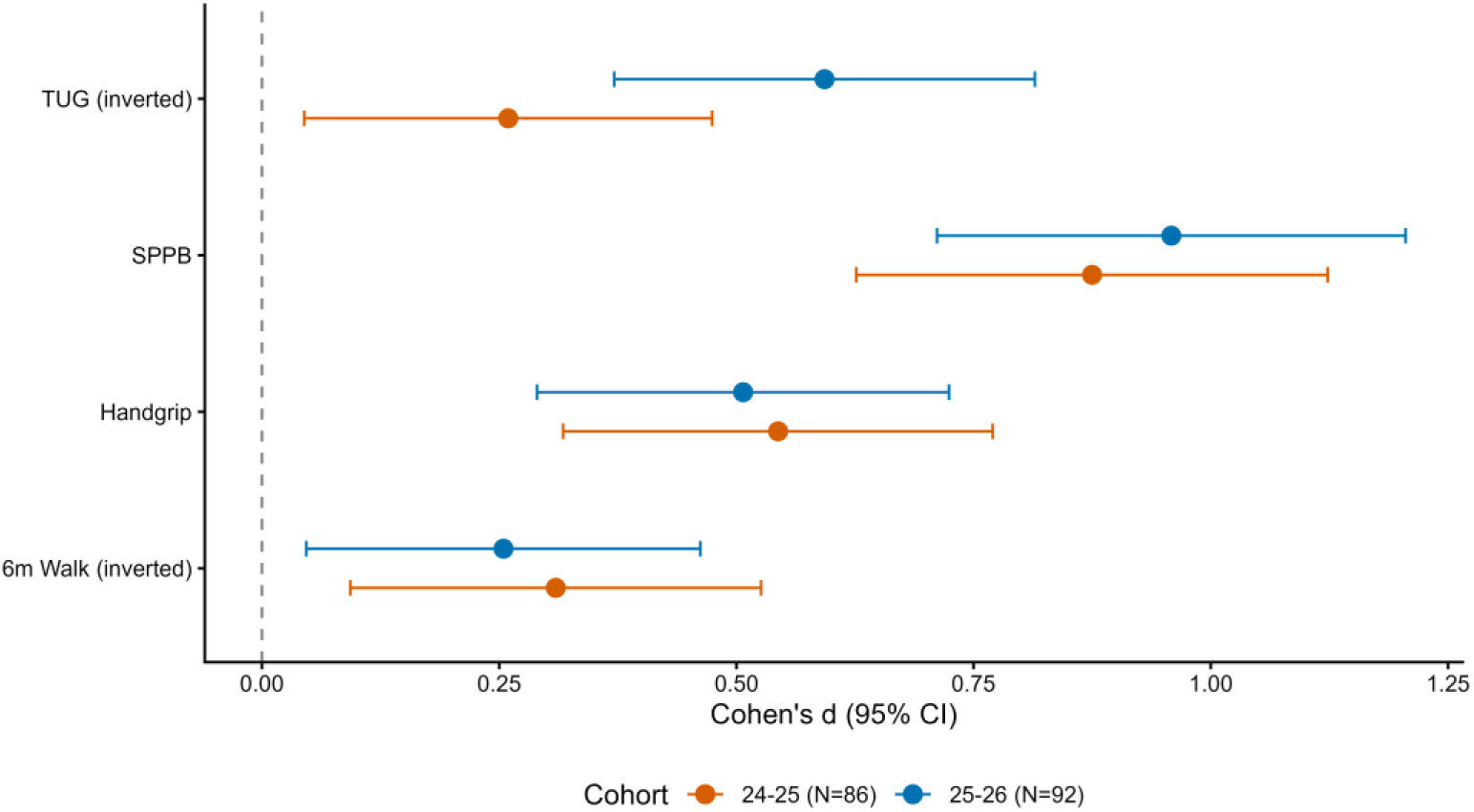
Replication forest plot: cohort-specific effect sizes (Cohen’s d, 95% CI) for the four shared core outcomes, 2024–2025 (n = 86) versus 2025–2026 (n = 92). TUG and 6-metre walk time are shown inverted (sign reversed) so that, as with SPPB and handgrip, a positive value represents improvement.

In the combined sample (n = 178), all four core outcomes improved significantly (Figure 6): SPPB (+1.16 points, dᵣ = 0.91), TUG (−1.24 s, dᵣ = −0.44), handgrip (+1.10 kg, dᵣ = 0.53), and 6-metre walk (−0.44 s, dᵣ = −0.28; all p < .001).

**Figure 6.**
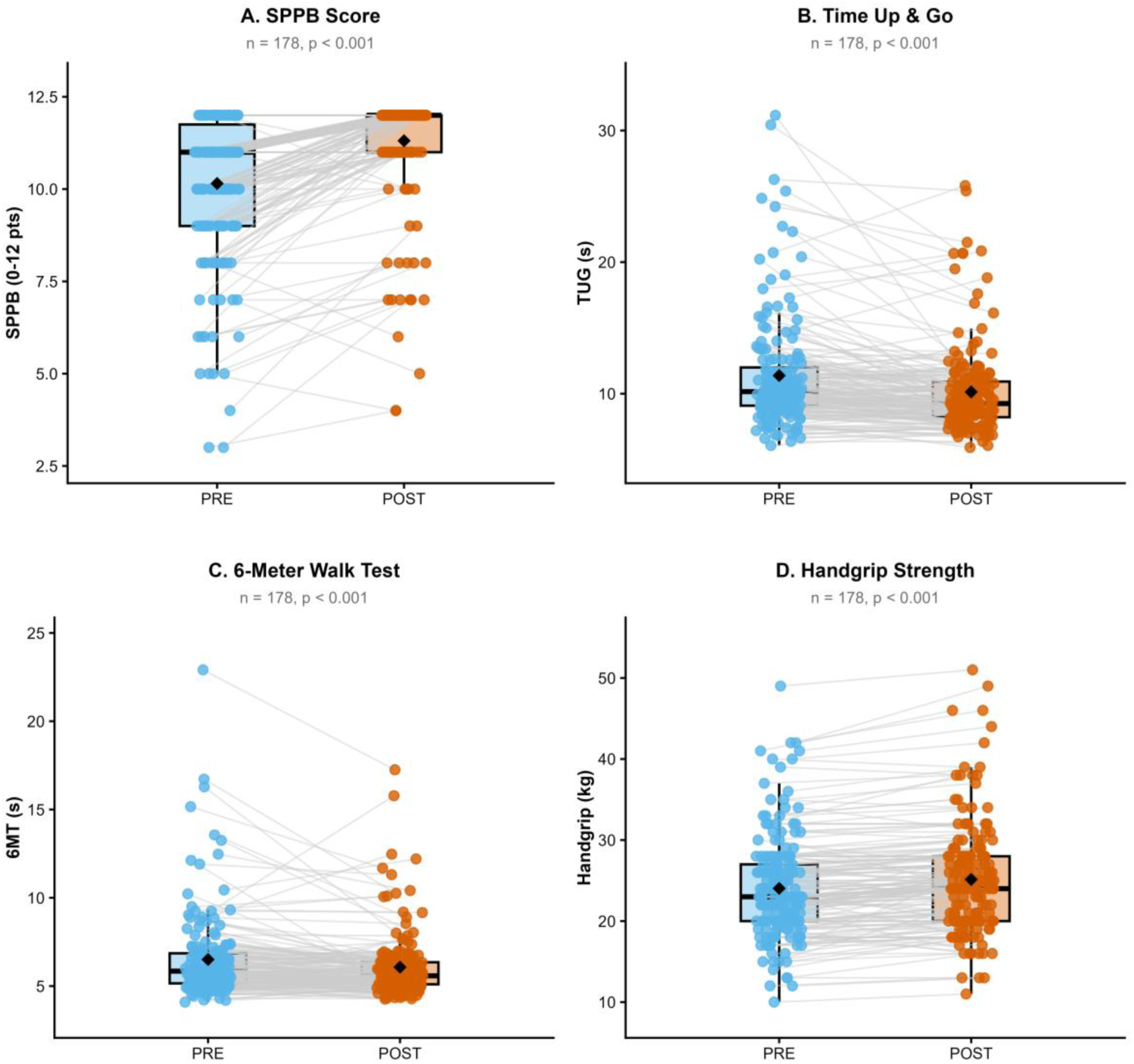
Pooled functional outcomes (paired PRE–POST), combined sample (n = 178). Black diamonds indicate the group mean.

Using the harmonised fall-risk algorithm, elevated fall risk fell from 19.7% to 11.2% in the pooled sample (McNemar χ² = 9.33, p = .002; Figure 7), with directionally consistent results in each cohort separately (2025–2026: 18.5% to 9.8%; 2024–2025: 20.9% to 12.8%).

**Figure 7.**
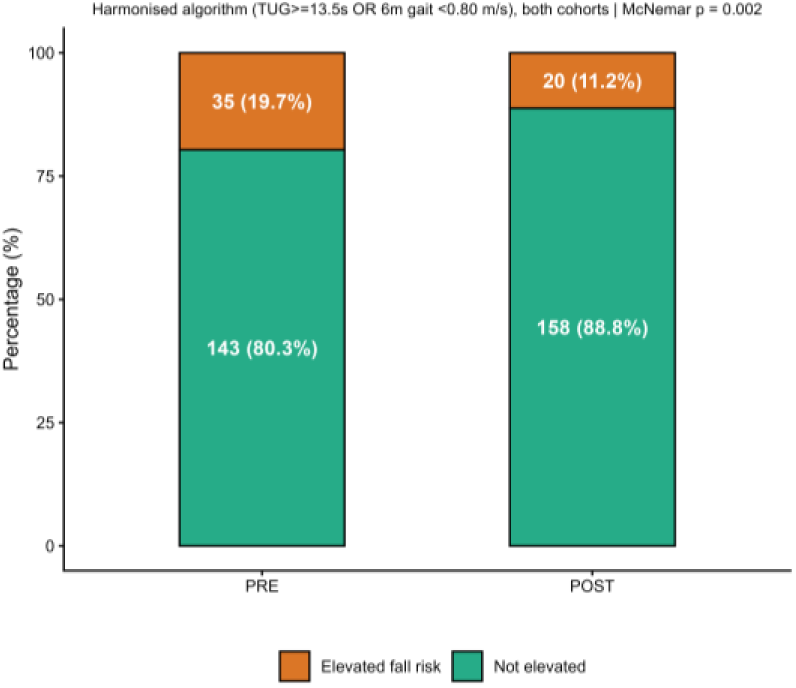
Fall-risk prevalence (TUG ≥13.5 s or gait speed <0.80 m/s) before and after the programme, pooled cohorts (n = 178).

SPPB MCID responder rates did not differ significantly between cohorts (72.8% vs. 62.8%; Fisher’s exact p = .198; Figure 8).

**Figure 8.**
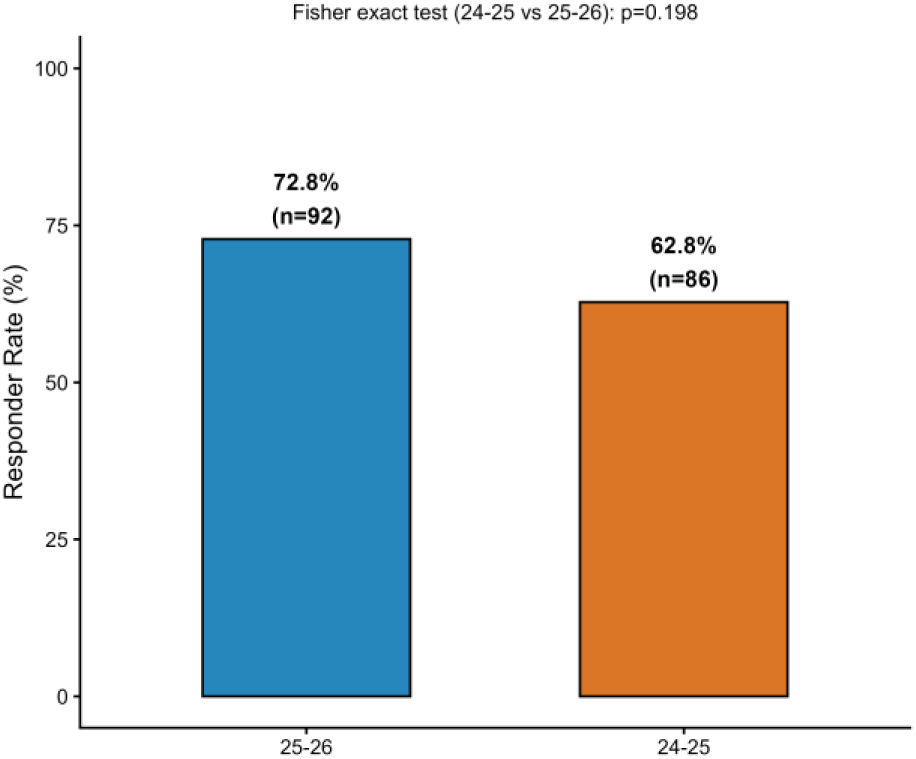
SPPB substantial-MCID responder rate by cohort, with Fisher’s exact test for the between-cohort comparison.

## 4. Discussion

### 4.1. SPPB, mobility and lower-limb performance

Across two consecutive cycles of this community multicomponent exercise programme, physical function improved between baseline and week 36. The clearest result is the SPPB change: large in the 2025–2026 cohort, and a mean increase of 1.16 points in the pooled sample of 178 participants. This magnitude aligns with the one-point threshold used as a clinically meaningful locomotor change in a responder analysis of a Vivifrail-based randomised trial, in which 67.4% of the exercise group reached that threshold versus 30.0% of controls (22); 72.8% of the 2025–2026 cohort reached the same threshold here. As a single-arm design, our study cannot rule out test practice, regression to the mean or seasonal variation; we therefore describe participants as having improved during programme participation rather than framing the programme as having caused the change. This pattern is consistent with the controlled evidence base: in the SPRINTT trial, a multicomponent intervention produced a 0.8- and 1.0-point SPPB difference at 24 and 36 months relative to an education control (23), and multicomponent exercise more broadly improves SPPB, TUG and strength in frail older adults (6,24).

The SPPB improvement is consistent with the programme content, and was accompanied by concordant gains in TUG, 5STS and both walk tests, handgrip strength (+1.10 kg pooled) and MMSE, though 4-metre gait and 5STS are themselves SPPB components and so only partly independent, and MMSE requires cautious interpretation given the uncontrolled design and possible ceiling effects. The larger TUG improvement in 2025–2026 than 2024–2025 is a genuine signal of between-cycle heterogeneity, plausibly reflecting TUG’s sensitivity to small differences in progression or test standardisation; the more defensible summary is that SPPB change was of similar magnitude with overlapping confidence intervals across cycles, rather than full replication of every outcome. Probable sarcopenia fell from 17.4% to 5.4%, best read as reclassification against the handgrip criterion used rather than confirmed reversal, since muscle mass was not measured; using an identical fall-risk algorithm in both cohorts, elevated fall risk fell from 19.7% to 11.2% in the pooled sample, consistent with meta-analytic evidence that multicomponent programmes reduce fall incidence, particularly at higher adherence (25).

### 4.2. Clinical response, adherence and dose-response

The 72.8% substantial-MCID responder rate offers a useful individual-level reading of the primary result. Descriptive similarity in responder proportions between the full analysis set and the per-protocol subset does not rule out adherence-related bias, since attendance reflects baseline capacity, motivation and support as well as dose received (26). In the adjusted analysis, attendance was not independently associated with SPPB change once baseline function, age and sex were accounted for, with no evidence of an attendance plateau or optimal threshold; the GO-OUT study similarly found attendance associated with endurance and gait-speed gains but not balance or strength, reflecting intervention-specific effects (27). Session-level measures of intensity and progression would likely be more informative than overall attendance for explaining this heterogeneity in future work.

### 4.3. Strengths and limitations

Strengths include implementation in a real municipal setting, two consecutive standardised cycles, high retention, and a functional battery spanning global performance, mobility, strength and clinical phenotypes; the multicohort analysis allows the consistency of core outcomes to be examined rather than relying on a single cycle. The main limitation is the absence of concurrent, randomised control, so pre-post change does not estimate a comparative causal effect; a pragmatic concurrent comparator and post-programme follow-up are needed to confirm durability. Inference is further limited by reduced power for subgroup interactions, particularly in men (n = 11), who accounted for only 12.0% of the sample and for whom findings should be generalised cautiously; by the absence of muscle-mass measurement to confirm sarcopenia; and by repeated use of tests susceptible to familiarisation.

### 4.4. Conclusions

Across two consecutive cycles of a 36-week community multicomponent exercise programme, participants showed a clinically interpretable improvement in SPPB, accompanied by favourable changes in mobility, gait and strength. The proportion reaching the substantial SPPB MCID adds a useful individual-level reading, but the single-arm design precludes attributing causality or confirming a dose-response relationship. The study contributes real-world implementation evidence and evidence of consistency across cycles for physical resilience in older adults; a concurrent comparator, post-programme follow-up and more precise measurement of dose and falls are the next steps needed to establish causal effectiveness and durability.

## Declaration of generative AI in scientific writing

During the preparation of this work, the author used Gemini 3.6 Thinking in order to refine the language and improve readability. After using this tool/service, the authors reviewed and edited the content as needed and take full responsibility for the content of the publication

## Funding

This research did not receive any specific grant from funding agencies in the public, commercial, or not-for-profit sectors. However, J.F.-O. was supported by a PhD fellowship from the Spanish Ministry of Science, Innovation and Universities (grant code: FPU22/03052).

## Data availability

The de-identified dataset and analysis code are available from the corresponding author upon reasonable request, subject to institutional data-sharing agreements.

## Supplementary Material

### Supplementary Methods

These formulae expand on the "Statistical Analysis" subsection of the main Methods. Cohen’s d for repeated measures (dᵣ) was calculated as dᵣ = Mean_diff / SD_diff, where SD_diff is the standard deviation of intra-individual paired differences. The 95% CI for dᵣ was approximated using SE_d = √(1/n + d²/2n). Mean and percentage change were calculated as Δ = Mean_POST−Mean_PRE and %Δ = (Δ/Mean_PRE) × 100.

The Tobit (censored-regression) sensitivity models for full-tandem hold time and 5STS completion time were fitted as accelerated failure-time models with a Gaussian distribution (survival::survreg), with time (PRE/POST), age and sex as covariates [16]. Because inability to complete a task does not necessarily correspond to a latent continuous time at the assumed censoring bound, these estimates are a bounded sensitivity check on the continuous-time components rather than a substitute for the protocol-scored SPPB outcome, which was used unmodified as the primary and secondary confirmatory analysis.

Restricted cubic splines (3 knots, evenly spaced across the attendance distribution) were fitted to the unadjusted attendance–ΔSPPB association, both within the 2025–2026 cohort and, adjusted for cohort, in the pooled sample. A likelihood-ratio test compared each spline model against the corresponding linear specification; an exploratory slope criterion (local slope <20% of the maximum slope) was used only to describe, not to formally test, the approximate point at which the fitted curve flattened.

### Supplementary Results

**Figure S1.**
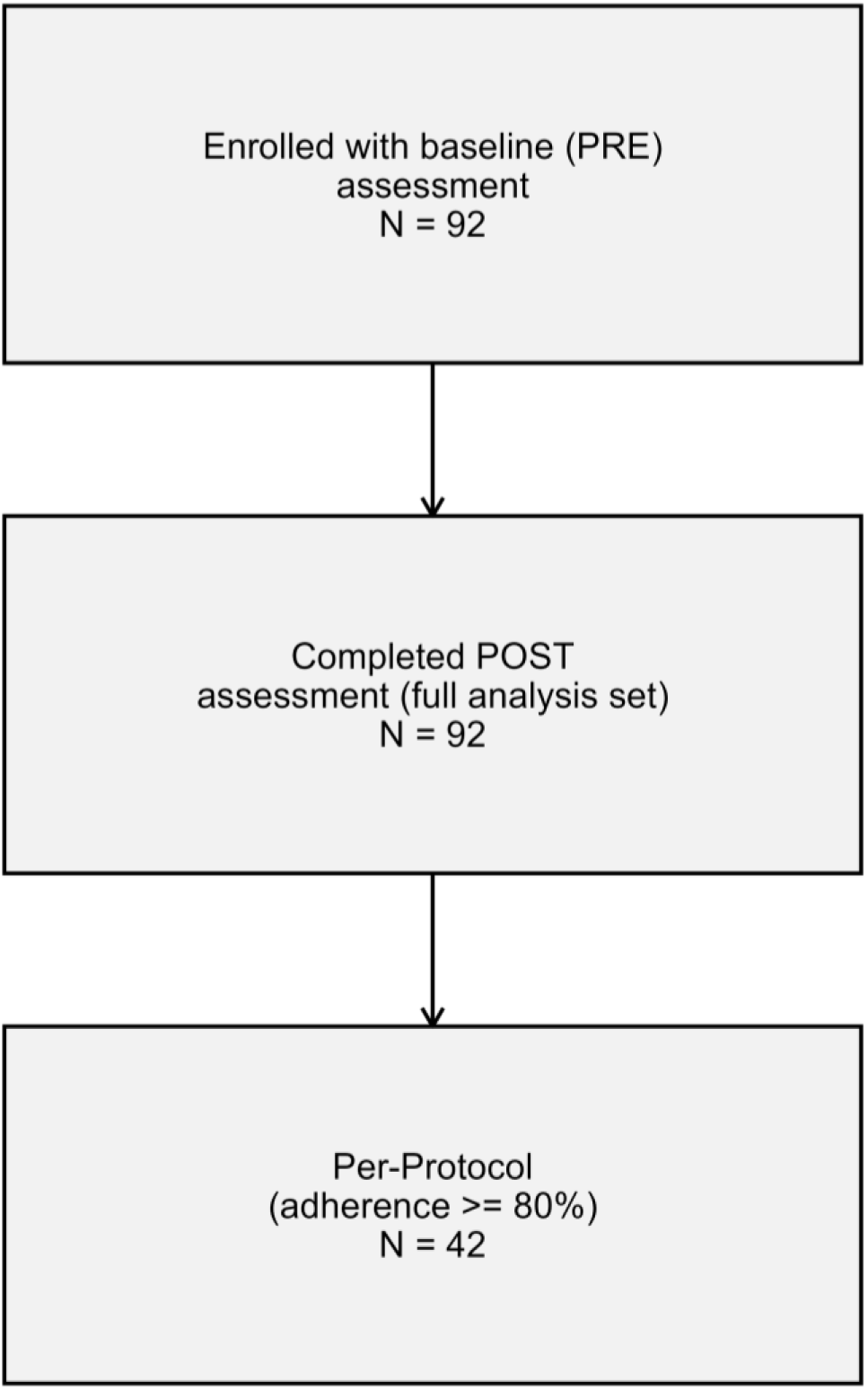
Flow diagram of participants through screening, enrolment, baseline functional assessment (PRE, n = 92), the 36-week multicomponent exercise programme, and post-intervention evaluation (POST, n = 92; 100% retention in the full analysis set). The per-protocol subset comprised participants achieving ≥80% programme attendance (n = 42).

**Table S1.** Functional outcomes, intention-to-treat/full analysis set (n = 92) versus per-protocol subset (≥80% attendance, n = 42).

| Outcome | Population | N | Mean change (SD) | $d_r$ | p (Wilcoxon) |
| --- | --- | --- | --- | --- | --- |
| **SPPB (points)** | Full analysis set | 92 | 1.33 (1.38) | 0.96 | <.001 |
| **SPPB (points)** | Per-protocol | 42 | 1.36 (1.32) | 1.03 | <.001 |
| **TUG (s)** | Full analysis set | 92 | -1.90 (3.21) | -0.59 | <.001 |
| **TUG (s)** | Per-protocol | 42 | -1.77 (2.52) | -0.70 | <.001 |
| **Handgrip (kg)** | Full analysis set | 92 | 1.00 (1.97) | 0.51 | <.001 |
| **Handgrip (kg)** | Per-protocol | 42 | 1.07 (1.88) | 0.57 | .002 |
| **6-m walk (s)** | Full analysis set | 92 | -0.45 (1.77) | -0.25 | .005 |
| **6-m walk (s)** | Per-protocol | 42 | -0.32 (1.25) | -0.25 | .149 |
| **MMSE (points)** | Full analysis set | 92 | 0.90 (1.76) | 0.51 | <.001 |
| **MMSE (points)** | Per-protocol | 42 | 1.40 (1.73) | 0.81 | <.001 |

**Table S2.**
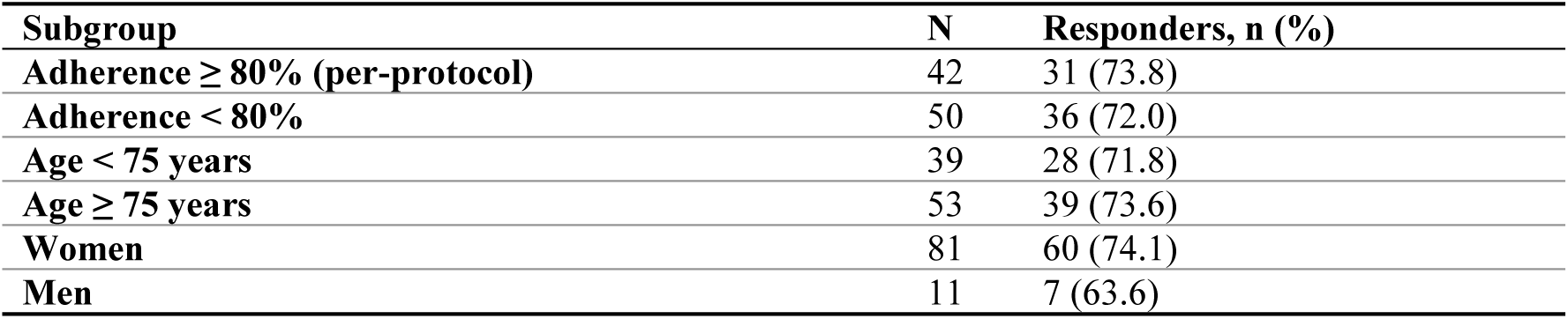
SPPB substantial-MCID responder rate (≥1.0 point) by subgroup, full analysis set.

Dose-response. Restricted cubic spline modelling of the unadjusted attendance-ΔSPPB association in the 2025–2026 cohort (Figure S2) did not show significant non-linearity relative to a linear specification (p = .095); the cohort-adjusted pooled spline model (n = 178; Figure S3) likewise approached but did not reach the conventional threshold (p = .061). Neither analysis provides sufficient evidence to characterise an attendance plateau or an optimal adherence threshold; both are reported as hypothesis-generating.

**Figure S2.**
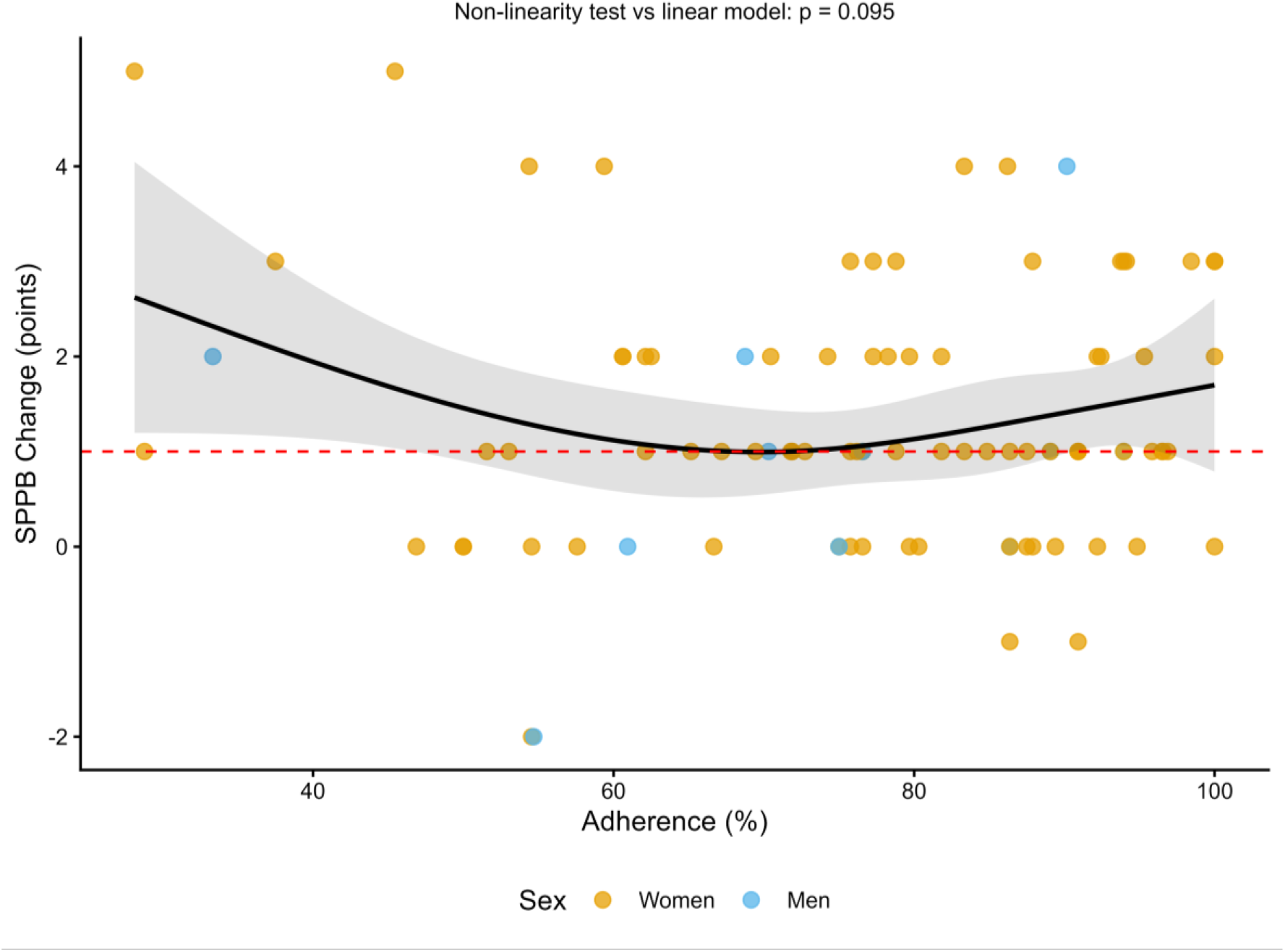
Dose-response between programme attendance and SPPB change, 2025–2026 cohort (n = 92), with an unadjusted restricted cubic spline (3 knots).

**Figure S3.**
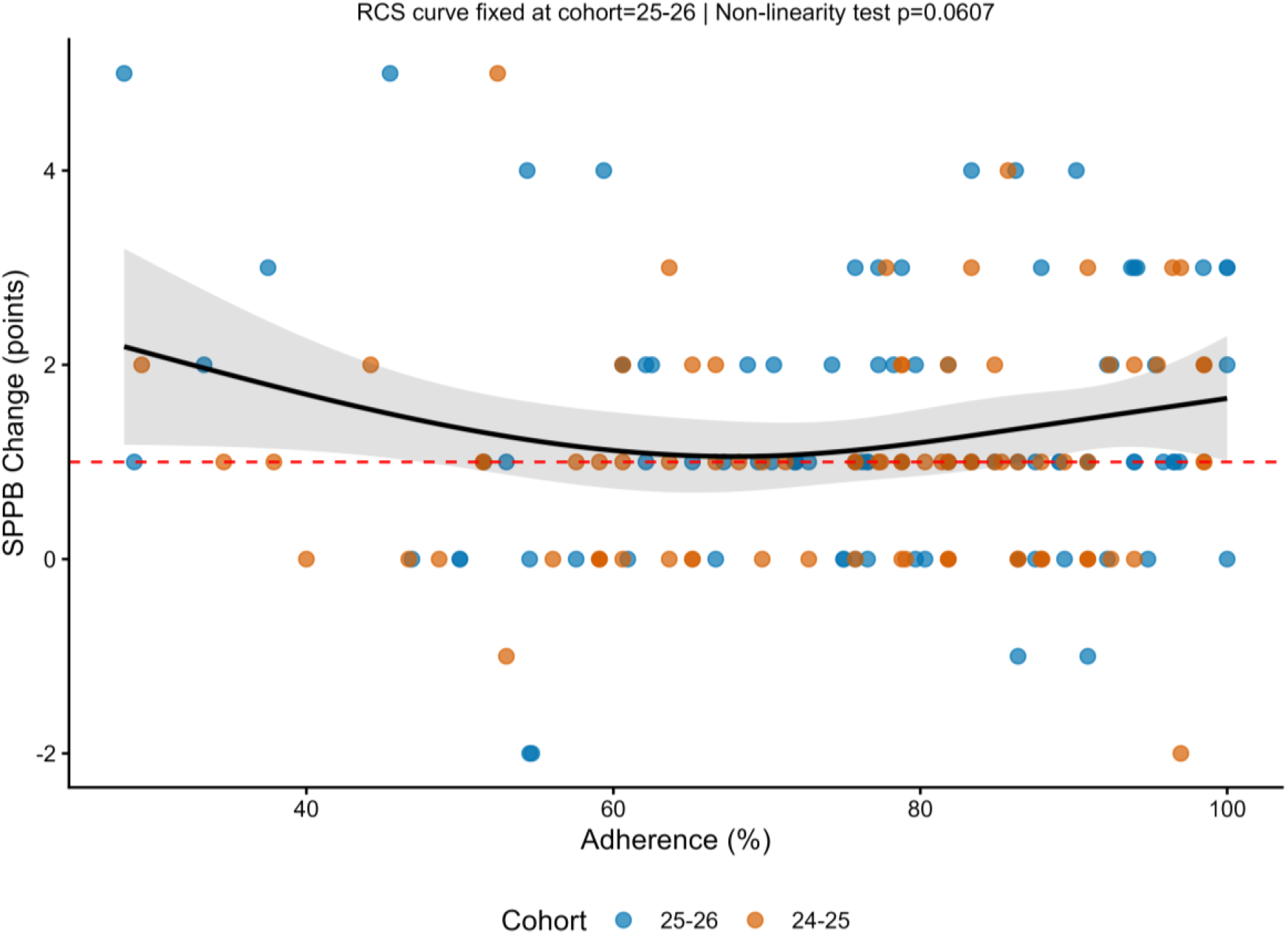
Dose-response between programme attendance and SPPB change, pooled cohorts (n = 178), cohort-adjusted restricted cubic spline.

Inter-domain correlations. Pairwise Pearson correlations among pre-to-post change scores (Figure S4) showed strong convergence among the timed mobility measures: Δ4-metre and Δ6-metre walk time (r = .96), ΔTUG and Δ6-metre walk time (r = .73), and ΔTUG and Δ4-metre walk time (r = .69), consistent with a shared gait-speed/mobility construct. Correlations between ΔSPPB and its own arithmetic components (Δ4-metre, Δ5STS) were not computed, as these are not independent by construction. Change in handgrip strength and MMSE correlated weakly with the timed mobility measures (|r| ≤ .20).

**Figure S4.**
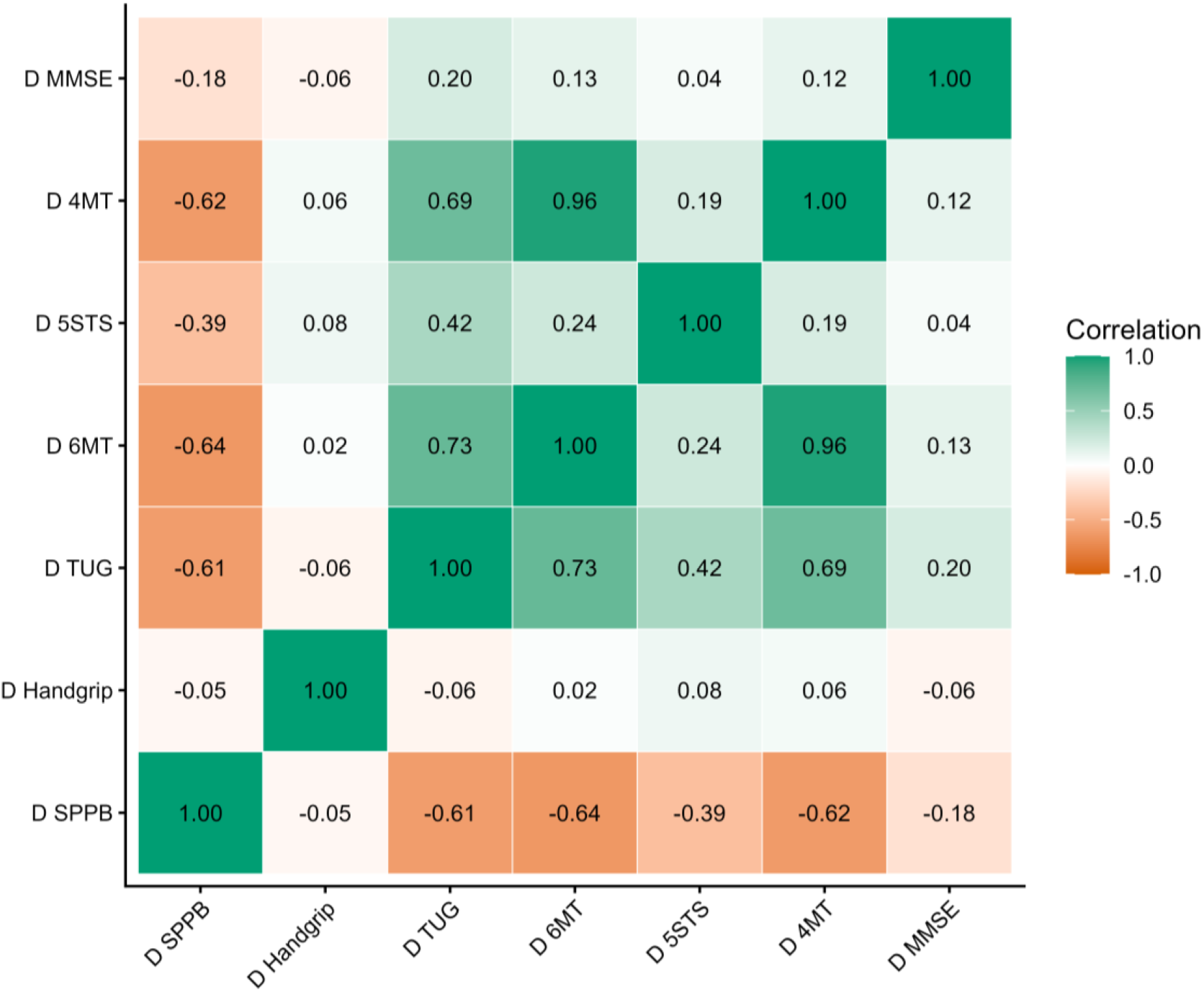
Correlation matrix (Pearson r) of pre-to-post change scores across functional and cognitive outcomes, 2025–2026 cohort (n = 92).

Effect sizes by domain. Figure S5 summarises the repeated-measures Cohen’s d (95% CI) for all seven functional and cognitive outcomes in the 2025–2026 cohort, ranked by magnitude; SPPB showed the largest effect, followed by TUG, MMSE, handgrip, 5STS, 4-metre walk and 6-metre walk.

**Figure S5.**
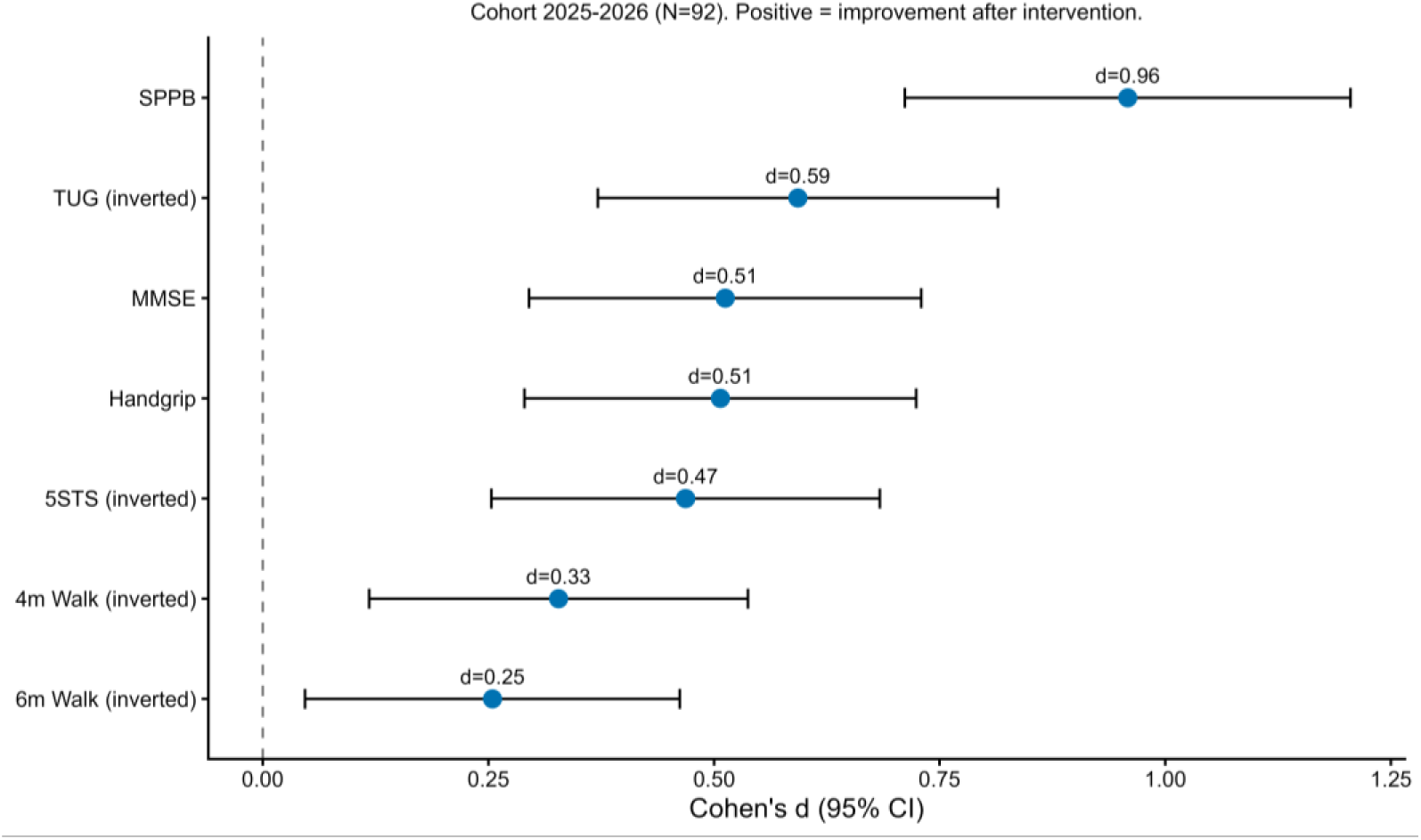
Repeated-measures effect sizes (Cohen’s d, 95% CI) for all functional and cognitive outcomes, 2025–2026 cohort (n = 92).

Responder rate by subgroup. Figure S6 displays the SPPB substantial-MCID responder proportion stratified by adherence, age group and sex (numerical values in Table S2).

**Figure S6.**
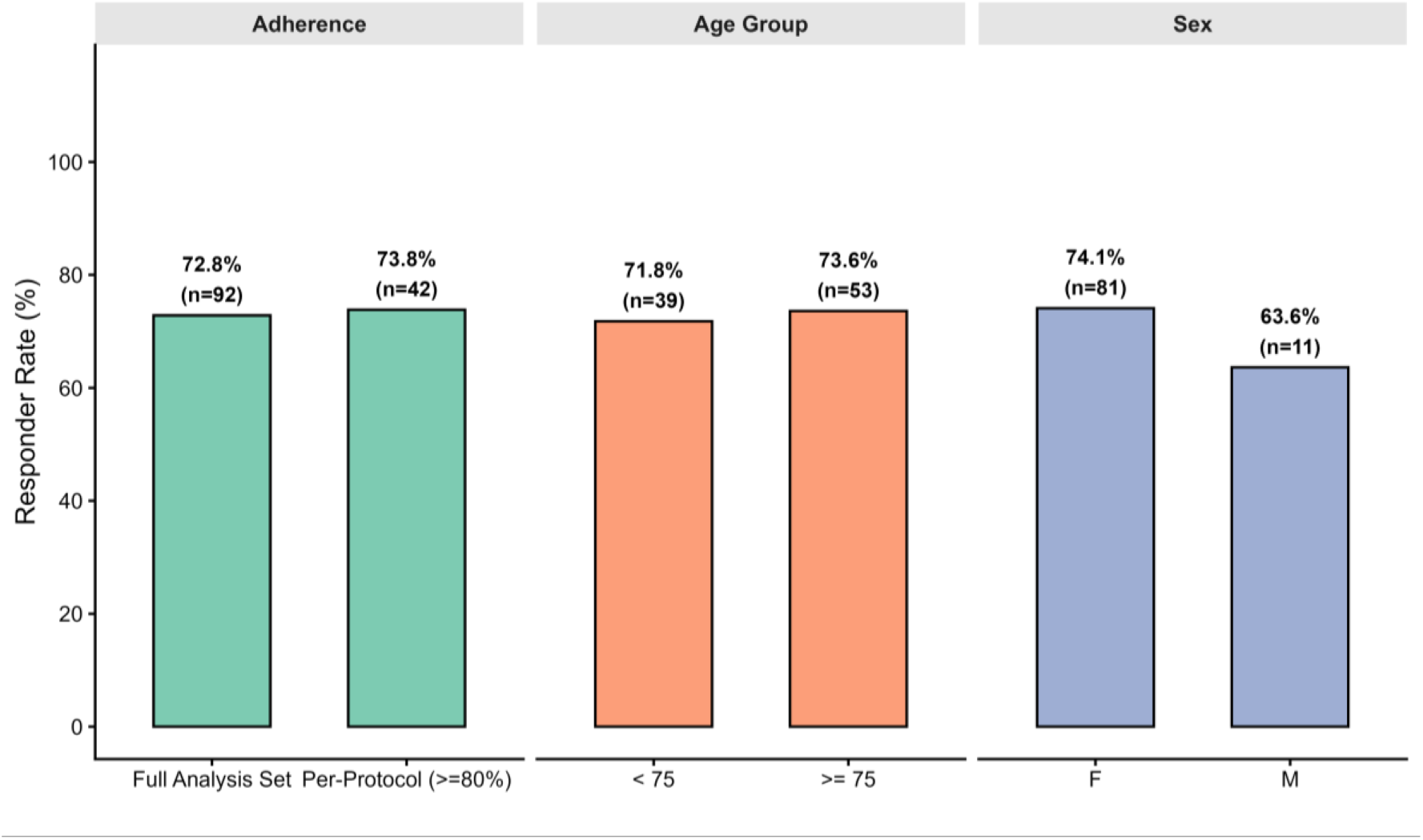
SPPB substantial-MCID responder rate (≥1.0 point) by adherence, age group and sex, full analysis set (n = 92).

## Notes

### Competing Interest Statement

The authors have declared no competing interest.

### Author Declarations

In accordance with national regulations for non-experimental healthcare service audits (Spanish Law 14/2007 on Biomedical Research), formal Institutional Review Board approval was not required, as the evaluation used secondary analysis of fully anonymised functional data collected during routine service delivery; no intervention was assigned for research purposes, and the study was accordingly not registered as a clinical trial

